# Meta-regression of COVID-19 prevalence/fatality on socioeconomic characteristics of data from top 50 U.S. large cities

**DOI:** 10.1101/2020.05.25.20112599

**Authors:** Hisato Takagi, Toshiki Kuno, Yujiro Yokoyama, Hiroki Ueyama, Takuya Matsushiro, Yosuke Hari, Tomo Ando

## Abstract

To screen potential risk and protective socioeconomic factors for Coronavirus disease 2019 (COVID-19) prevalence and fatality, meta-regression of data from top 50 U.S. large-population cities was performed. The population estimate (in 2019) of each country to which the city belongs was abstracted from the “County Population Totals: 2010-2019.” From the “Johns Hopkins Coronavirus Resource Center,” the cumulative number of confirmed cases and deaths of COVID-19 in each country was obtained on May 22, 2020. Socioeconomic characteristics of each country were extracted from the “2014-2018 American Community Survey (ACS) 5-Year Data Profile” and “Small Area Income and Poverty Estimates (SAIPE) Program (for 2018).” Radom-effects meta-regression was performed using OpenMetaAnalyst (http://www.cebm.brown.edu/openmeta/index.html). A coefficient (slope of the meta-regression line) for COVID-19 prevalence was significantly negative for male sex, education attainment, computer and Internet use, and private health insurance. Whereas, the coefficient was significantly positive for black race, never matrimony, unemployment, and poverty. In the multivariable model, the coefficient was significantly negative for male sex (P = 0.036) and computer use (P = 0.024), and significantly positive for never matrimony (P < 0.001). A coefficient for COVID-19 fatality was significantly negative for no health insurance, and significantly positive for elderly, unemployment, and public coverage. In the multivariable model, the coefficient was significantly positive for only elderly (P = 0.002). In conclusion, a number of socioeconomic factors, e.g. male sex (negatively for prevalence), elderly (positively for fatality), never matrimony (positively for prevalence), and computer use (negatively for prevalence) may be associated with COVID-19.

## Introduction

Reuters reported “The coronavirus mortality rate among some of the poorest Catalans is five times higher than among the wealthiest residents of the Spanish region, a study showed, in the latest evidence of how COVID-19 [Coronavirus disease 2019] hits the needy hardest” on 22 May, 2020.^1^ It has been suggested that outcomes of pandemic influenza are associated with socioeconomic status.^2^ Socioeconomic characteristics also may affect prevalence and case fatality of COVID-19. To screen potential risk and protective socioeconomic factors for COVID-19 prevalence and fatality, meta-regression of data from top 50 U.S. large cities was performed.

## Methods

From the “Population and Housing Unit Estimates (https://www.census.gov/programs-surveys/popest.html),” top 50 U.S. large-population cities (in 2019) were selected. The population estimate (in 2019) of each country to which the city belongs was abstracted from the “County Population Totals: 2010-2019 (https://www.census.gov/data/tables/time-series/demo/popest/2010s-counties-total.html).” From the “Johns Hopkins Coronavirus Resource Center (https://coronavirus.jhu.edu/us-map),” the cumulative number of confirmed cases and deaths of COVID-19 in each country was obtained on May 22, 2020. Socioeconomic characteristics of each country were extracted from the “2014-2018 American Community Survey (ACS) 5-Year Data Profile (https://www.census.gov/acs/www/data/data-tables-and-tools/data-profiles/)” and “Small Area Income and Poverty Estimates (SAIPE) Program (for 2018) (https://www.census.gov/programs-surveys/saipe.html),” which included male (%), 65 years and over (%), black/African American (%), Hispanic/Latino (%), never married in population 15 years and over (%), high school graduate or higher in population 25 years and over (%), households with a computer (%) and broadband Internet subscription (%), unemployment rate (%), median (dollars) and mean (dollars) household income, civilian noninstitutionalized population with private health insurance (%), public coverage (%), and no health insurance coverage (%), and poverty rate (%) (Table 1). Radom-effects meta-regression was performed using OpenMetaAnalyst (http://www.cebm.brown.edu/openmeta/index.html). A meta-regression graph depicted the COVID-19 prevalence or fatality (plotted as the logarithm-transformed prevalence or fatality on the y-axis) as a function of a given factor (plotted as a socioeconomic characteristic on the x-axis). Covariates with a significantly (P < 0.05) positive or negative coefficient in the univariable model were together entered into the multivariable model.

**Table 1.** Demographic/socioeconomic characteristics and COVID-19 prevalence/fatality in the country to which the city belongs.

| Geographic Area |  |  | Country |  |  |  |  |  |  |
| --- | --- | --- | --- | --- | --- | --- | --- | --- | --- |
| City | State | Population estimate (N) | Rank |  | Population estimate (N) | Confirmed cases |  | Deaths |  |
|  |  |  |  |  |  | N | % | N | % |
| New York city | New York | 8,336,817 | 1 | Bronx | 1,418,207 | 43,918 | 3.10 | 3,459 | 7.88 |
|  |  |  |  | Kings | 2,559,903 | 53,137 | 2.08 | 4,969 | 9.35 |
|  |  |  |  | New York | 1,628,706 | 24,101 | 1.48 | 2,221 | 9.22 |
|  |  |  |  | Queens | 2,253,858 | 59,615 | 2.65 | 4,887 | 8.20 |
|  |  |  |  | Richmond | 476,143 | 13,080 | 2.75 | 785 | 6.00 |
| Los Angeles city | California | 3,979,576 | 2 | Los Angeles | 10,039,107 | 43,070 | 0.43 | 2,053 | 4.77 |
| Chicago city | Illinois | 2,693,976 | 3 | Cook | 5,150,233 | 68,949 | 1.34 | 3,187 | 4.62 |
| Houston city | Texas | 2,320,268 | 4 | Harris | 4,713,325 | 10,526 | 0.22 | 217 | 2.06 |
| Phoenix city | Arizona | 1,680,992 | 5 | Maricopa | 4,485,414 | 7,950 | 0.18 | 368 | 4.63 |
| Philadelphia city | Pennsylvania | 1,584,064 | 6 | Philadelphia | 1,584,064 | 21,009 | 1.33 | 1,221 | 5.81 |
| San Antonio city | Texas | 1,547,253 | 7 | Bexar | 2,003,554 | 2,371 | 0.12 | 64 | 2.70 |
| San Diego city | California | 1,423,851 | 8 | San Diego | 3,338,330 | 6,315 | 0.19 | 241 | 3.82 |
| Dallas city | Texas | 1,343,573 | 9 | Dallas | 2,635,516 | 8,477 | 0.32 | 207 | 2.44 |
| San Jose city | California | 1,021,795 | 10 | Santa Clara | 1,927,852 | 2,492 | 0.13 | 138 | 5.54 |
| Austin city | Texas | 978,908 | 11 | Travis | 1,273,954 | 2,644 | 0.21 | 82 | 3.10 |
| Jacksonville city | Florida | 911,507 | 12 | Duval | 957,755 | 1,376 | 0.14 | 41 | 2.98 |
| Fort Worth city | Texas | 909,585 | 13 | Tarrant | 2,102,515 | 4,711 | 0.22 | 132 | 2.80 |
| Columbus city | Ohio | 898,553 | 14 | Franklin | 1,316,756 | 4,996 | 0.38 | 215 | 4.30 |
| Charlotte city | North Carolina | 885,708 | 15 | Mecklenburg | 1,110,356 | 3,001 | 0.27 | 73 | 2.43 |
| San Francisco city | California | 881,549 | 16 | San Francisco | 881,549 | 2,320 | 0.26 | 40 | 1.72 |
| Indianapolis city (balance) | Indiana | 876,384 | 17 | Marion | 964,582 | 8,928 | 0.93 | 528 | 5.91 |
| Seattle city | Washington | 753,675 | 18 | King | 2,252,782 | 7,669 | 0.34 | 542 | 7.07 |
| Denver city | Colorado | 727,211 | 19 | Denver | 727,211 | 5,056 | 0.70 | 299 | 5.91 |
| Washington city | District of Columbia | 705,749 | 20 | District of Columbia | 705,749 | 7,893 | 1.12 | 418 | 5.30 |
| Boston city | Massachusetts | 692,600 | 21 | Suffolk | 803,907 | 17,180 | 2.14 | 818 | 4.76 |
| El Paso city | Texas | 681,728 | 22 | El Paso | 839,238 | 2,046 | 0.24 | 57 | 2.79 |
| Nashville-Davidson metropolitan government (balance) | Tennessee | 670,820 | 23 | Davidson | 694,144 | 4,396 | 0.63 | 48 | 1.09 |
| Detroit city | Michigan | 670,031 | 24 | Wayne | 1,749,343 | 19,602 | 1.12 | 2,323 | 11.85 |
| Oklahoma City city | Oklahoma | 655,057 | 25 | Oklahoma | 797,434 | 1,179 | 0.15 | 55 | 4.66 |
| Portland city | Oregon | 654,741 | 26 | Multnomah | 812,855 | 1,037 | 0.13 | 58 | 5.59 |
| Las Vegas city | Nevada | 651,319 | 27 | Clark | 2,266,715 | 5,815 | 0.26 | 322 | 5.54 |
| Memphis city | Tennessee | 651,073 | 28 | Shelby | 937,166 | 4,127 | 0.44 | 92 | 2.23 |
| Louisville/Jefferson<br>County metro<br>government (balance) | Kentucky | 617,638 | 29 | Jefferson | 766,757 | 2,021 | 0.26 | 141 | 6.98 |
| Baltimore city | Maryland | 593,490 | 30 | Baltimore city | 593,490 | 4,492 | 0.76 | 220 | 4.90 |
| Milwaukee city | Wisconsin | 590,157 | 31 | Milwaukee | 945,726 | 5,735 | 0.61 | 270 | 4.71 |
| Albuquerque city | New Mexico | 560,513 | 32 | Bernalillo | 679,121 | 1,313 | 0.19 | 66 | 5.03 |
| Tucson city | Arizona | 548,073 | 33 | Pima | 1,047,279 | 1,974 | 0.19 | 174 | 8.81 |
| Fresno city | California | 531,576 | 34 | Fresno | 999,101 | 1,372 | 0.14 | 22 | 1.60 |
| Mesa city | Arizona | 518,012 | 35 | (Maricopa) | (4,485,414) | – | – | – | – |
| Sacramento city | California | 513,624 | 36 | Sacramento | 1,552,058 | 1,272 | 0.08 | 56 | 4.40 |
| Atlanta city | Georgia | 506,811 | 37 | Fulton | 1,063,937 | 3,931 | 0.37 | 196 | 4.99 |
| Kansas City city | Missouri | 495,327 | 38 | Jackson | 703,011 | 485 | 0.07 | 16 | 3.30 |
| Colorado Springs city | Colorado | 478,221 | 39 | El Paso | 720,403 | 1,460 | 0.20 | 88 | 6.03 |
| Omaha city | Nebraska | 478,192 | 40 | Douglas | 571,327 | 2,881 | 0.50 | 23 | 0.80 |
| Raleigh city | North Carolina | 474,069 | 41 | Wake | 1,111,761 | 1,393 | 0.13 | 32 | 2.30 |
| Miami city | Florida | 467,963 | 42 | Miami-Dade | 2,716,940 | 16,522 | 0.61 | 614 | 3.72 |
| Long Beach city | California | 462,628 | 43 | (Los Angeles) | (10,039,107) | – | – | – | – |
| Virginia Beach city | Virginia | 449,974 | 44 | Virginia<br>Beach city | 449,974 | 591 | 0.13 | 19 | 3.21 |
| Oakland city | California | 433,031 | 45 | Alameda | 1,671,329 | 2,708 | 0.16 | 90 | 3.32 |
| Minneapolis city | Minnesota | 429,606 | 46 | Hennepin | 1,265,843 | 6,350 | 0.50 | 518 | 8.16 |
| Tulsa city | Oklahoma | 401,190 | 47 | Tulsa | 651,552 | 890 | 0.14 | 41 | 4.61 |
| Tampa city | Florida | 399,700 | 48 | Hillsborough | 1,471,968 | 1,790 | 0.12 | 70 | 3.91 |
| Arlington city | Texas | 398,854 | 49 | (Tarrant) | -2,102,515 | – | – | – | – |
| New Orleans city | Louisiana | 390,144 | 50 | Orleans Parish | 390,144 | 6,944 | 1.78 | 500 | 7.20 |

Table 1. *Continued.*
| Country | Male (%) | 65 years and over (%) | Black/African American (%) | Hispanic/Latino (%) | Never married in population 15 years and over (%) | High school graduate or higher in population 25 years and over (%) | Households |  |
| --- | --- | --- | --- | --- | --- | --- | --- | --- |
|  |  |  |  |  |  |  | With a computer (%) | With a broadband Internet subscription (%) |
| Bronx | 47.1 | 12.1 | 34.1 | 55.9 | 48.5 | 72 | 84.3 | 72.5 |
| Kings | 47.4 | 13.2 | 32.6 | 19.2 | 43.8 | 81.6 | 85.8 | 77.6 |
| New York | 47.3 | 15.8 | 14.8 | 26 | 49.5 | 87 | 90.1 | 83.3 |
| Queens | 48.5 | 14.8 | 18.3 | 28 | 37.8 | 81.5 | 88.8 | 81.9 |
| Richmond | 48.4 | 15.5 | 10.2 | 18.3 | 33.0 | 88.5 | 88.3 | 81.1 |
| Los Angeles | 49.3 | 12.9 | 8.2 | 48.5 | 41.8 | 78.7 | 90.4 | 82.1 |
| Cook | 49.1 | 14.8 | 14.2 | 17.6 | 41.6 | 86.7 | 87.6 | 78.9 |
| Harris | 49.7 | 9.8 | 19 | 42.6 | 36.3 | 80.9 | 89.9 | 81 |
| Maricopa | 49.5 | 14.5 | 5.5 | 30.8 | 34.3 | 87.4 | 91.5 | 84.1 |
| Philadelphia | 47.3 | 13.2 | 42.3 | 14.5 | 50.9 | 83.9 | 84.1 | 73.7 |
| Bexar | 49.3 | 11.6 | 7.6 | 60 | 36.8 | 83.8 | 89.3 | 78.9 |
| San Diego | 50.3 | 13.3 | 5 | 33.5 | 35.8 | 87.1 | 93.9 | 88.8 |
| Dallas | 49.3 | 10.2 | 22.5 | 39.9 | 37.2 | 78.7 | 88.7 | 78.5 |
| Santa Clara | 50.5 | 12.8 | 2.5 | 25.8 | 33.5 | 88.1 | 95.3 | 90.7 |
| Travis | 50.5 | 9.2 | 8.3 | 33.9 | 40.0 | 89.1 | 94.2 | 86.5 |
| Duval | 48.5 | 13.4 | 29.6 | 9.3 | 34.7 | 89.5 | 89.4 | 81.2 |
| Tarrant | 48.9 | 10.7 | 16.2 | 28.5 | 32.9 | 85.6 | 93.1 | 84.3 |
| Franklin | 48.8 | 11.4 | 22.2 | 5.4 | 39.4 | 91 | 91.6 | 85.3 |
| Mecklenburg | 48.1 | 10.6 | 31.3 | 13 | 37.6 | 90.1 | 93.4 | 86.5 |
| San Francisco | 51 | 15.1 | 5.2 | 15.2 | 45.9 | 88.5 | 91.9 | 86 |
| Marion | 48.2 | 12 | 27.7 | 10.2 | 41.6 | 85.7 | 85.5 | 76.5 |
| King | 50.1 | 12.7 | 6.3 | 9.6 | 34.5 | 93 | 94.9 | 90.2 |
| Denver | 50.1 | 11.4 | 9.4 | 30.3 | 42.5 | 87.1 | 91.8 | 83.7 |
| District of Columbia | 47.5 | 11.9 | 46.9 | 10.9 | 56.3 | 90.6 | 89.8 | 80 |
| Suffolk | 48.4 | 11.5 | 22.4 | 22.7 | 53.7 | 85.5 | 89.1 | 82.8 |
| El Paso | 49.1 | 11.6 | 3.4 | 82.4 | 32.8 | 83.2 | 89.2 | 79.3 |
| Davidson | 48.2 | 11.7 | 27.4 | 10.2 | 40.0 | 88.6 | 90.8 | 82.5 |
| Wayne | 48.1 | 14.7 | 38.9 | 5.9 | 41.2 | 85.9 | 85.6 | 72.1 |
| Oklahoma | 48.9 | 13 | 15.1 | 17.1 | 32.7 | 87.2 | 88.8 | 79.6 |
| Multnomah | 49.5 | 12.6 | 5.4 | 11.4 | 39.2 | 91.3 | 93 | 86.1 |
| Clark | 50 | 14.1 | 11.5 | 30.9 | 34.8 | 85.7 | 91.4 | 81 |
| Shelby | 47.6 | 12.7 | 53.5 | 6.2 | 41.6 | 88.2 | 83.2 | 72.6 |
| Jefferson | 48.3 | 15.3 | 21.5 | 5.2 | 34.6 | 90 | 87.3 | 80.5 |
| Baltimore city | 47 | 13.2 | 62.5 | 5.1 | 52.6 | 84.9 | 84 | 72.3 |
| Milwaukee | 48.4 | 12.9 | 26.4 | 14.7 | 44.8 | 87.8 | 85.2 | 76.3 |
| Bernalillo | 49 | 15.2 | 2.8 | 49.8 | 36.0 | 88.8 | 88.4 | 77.7 |
| Pima | 49.2 | 18.7 | 3.8 | 37 | 34.4 | 88.3 | 91 | 83.1 |
| Fresno | 49.9 | 11.7 | 4.8 | 52.7 | 38.8 | 75.3 | 87.7 | 77.9 |
| (Maricopa) | – | – | – | – | – | – | – | – |
| Sacramento | 48.9 | 13.4 | 9.9 | 23 | 35.2 | 87.4 | 93.2 | 85.6 |
| Fulton | 48.4 | 11.1 | 44.1 | 7.3 | 44.5 | 92.1 | 91.6 | 83.1 |
| Jackson | 48.4 | 14.3 | 23.6 | 8.9 | 36.1 | 90.3 | 87.3 | 78.7 |
| El Paso | 50.5 | 12.1 | 6.3 | 16.8 | 29.7 | 93.7 | 93.8 | 89.3 |
| Douglas | 49.3 | 12.3 | 11.1 | 12.4 | 34.2 | 90 | 89 | 82.8 |
| Wake | 48.6 | 10.9 | 20.3 | 10.1 | 33.1 | 92.7 | 95.5 | 89.7 |
| Miami-Dade | 48.6 | 15.6 | 17.7 | 68 | 36.8 | 81.5 | 87.7 | 75.1 |
| (Los Angeles) | – | – | – | – | – | – | – | – |
| Virginia Beach city | 49.2 | 13.2 | 19 | 8 | 30.7 | 93.2 | 94.4 | 87.8 |
| Alameda | 49.1 | 13.1 | 10.8 | 22.5 | 36.8 | 88 | 93.6 | 87.5 |
| Hennepin | 49.4 | 13.3 | 13.1 | 6.9 | 37.0 | 93.2 | 92.6 | 85.3 |
| Tulsa | 48.8 | 13.6 | 10.1 | 12.4 | 30.5 | 89.2 | 89.8 | 81.8 |
| Hillsborough | 48.8 | 13.8 | 16.6 | 28 | 34.8 | 88.4 | 92 | 85 |
| (Tarrant) | – | – | – | – | – | – | – | – |
| Orleans Parish | 47.6 | 13.5 | 59.7 | 5.5 | 49.2 | 86.2 | 81.8 | 69.6 |

Table 1. *Continued.*
| Country | Unemployment<br>rate (%) | Household income |  | Civilian noninstitutionalized population |  |  | Poverty<br>rate (%) |
| --- | --- | --- | --- | --- | --- | --- | --- |
|  |  | Median<br>(dollars) | Mean<br>(dollars) | With private health<br>insurance (%) | With public<br>coverage (%) | No health insurance<br>coverage (%) |  |
| Bronx | 10.5 | 38,085 | 56,328 | 44.7 | 55.7 | 9.7 | 27.3 |
| Kings | 7 | 56,015 | 85,910 | 54.5 | 45.1 | 8.2 | 18.9 |
| New York | 5.7 | 82,459 | 152,002 | 68.8 | 34 | 5.8 | 15.6 |
| Queens | 6.2 | 64,987 | 85,330 | 43 | 47.7 | 15.1 | 11.6 |
| Richmond | 4.9 | 79,267 | 98,841 | 85.9 | 9.8 | 5.9 | 11.7 |
| Los Angeles | 6.8 | 64,251 | 94,484 | 57.9 | 38 | 10.8 | 14.2 |
| Cook | 7.8 | 62,088 | 91,216 | 64.4 | 34.8 | 9.5 | 13.8 |
| Harris | 6.1 | 60,146 | 89,270 | 57.7 | 28.3 | 20.2 | 16.5 |
| Maricopa | 5.5 | 61,606 | 84,884 | 65.1 | 33.9 | 11.1 | 12.3 |
| Philadelphia | 10.2 | 43,744 | 65,090 | 57 | 44.1 | 9.1 | 24.3 |
| Bexar | 5.8 | 55,456 | 76,006 | 77.8 | 7.2 | 18.6 | 17.2 |
| San Diego | 6.4 | 74,855 | 101,302 | 68.2 | 33 | 8.7 | 11.5 |
| Dallas | 5.2 | 56,854 | 85,064 | 55.6 | 29.6 | 21 | 14.2 |
| Santa Clara | 4.8 | 116,178 | 154,183 | 76.7 | 26.8 | 4.8 | 7.3 |
| Travis | 4 | 71,767 | 103,148 | 72.7 | 21.3 | 13.5 | 12 |
| Duval | 6.6 | 53,473 | 74,331 | 64.8 | 34 | 12 | 14.5 |
| Tarrant | 5 | 64,874 | 89,207 | 65.2 | 26 | 16.3 | 12.1 |
| Franklin | 5.8 | 54,533 | 74,109 | 69.5 | 36.6 | 6.5 | 15.5 |
| Mecklenburg | 5.8 | 64,312 | 93,951 | 70.4 | 25.6 | 11.9 | 11.7 |
| San Francisco | 4.7 | 104,552 | 149,265 | 75.2 | 29.1 | 4.4 | 10.1 |
| Marion | 7.1 | 46,692 | 65,991 | 79 | 9.8 | 13.5 | 17.2 |
| King | 4.5 | 89,418 | 120,828 | 77.3 | 26.6 | 5.7 | 9.2 |
| Denver | 4 | 63,793 | 93,650 | 64.6 | 33.2 | 10.3 | 11.7 |
| District of<br>Columbia | 7.4 | 82,604 | 121,698 | 69.6 | 36 | 4 | 16.1 |
| Suffolk | 7 | 64,582 | 97,859 | 64.7 | 39.2 | 4 | 17.5 |
| El Paso | 6.6 | 44,597 | 60,632 | 51.7 | 34.2 | 20.7 | 20.5 |
| Davidson | 4.6 | 56,507 | 81,577 | 66.5 | 29.9 | 12.6 | 15.4 |
| Wayne | 10.2 | 45,321 | 65,173 | 59.8 | 46.4 | 7.2 | 21.7 |
| Oklahoma | 4.9 | 52,855 | 76,925 | 64.3 | 32.7 | 14.4 | 16.9 |
| Multnomah | 5.6 | 64,337 | 88,399 | 69 | 34.2 | 7.1 | 12 |
| Clark | 7.2 | 56,802 | 76,812 | 64.3 | 32.8 | 12.5 | 14 |
| Shelby | 7.7 | 49,782 | 73,887 | 62.6 | 35.9 | 12.1 | 21.7 |
| Jefferson | 5.9 | 54,357 | 76,517 | 69.8 | 38 | 5.5 | 15.4 |
| Baltimore city | 9.1 | 48,840 | 71,259 | 57.7 | 45.5 | 7.2 | 18.9 |
| Milwaukee | 6.2 | 48,742 | 67,364 | 63.1 | 39.3 | 7.8 | 19.1 |
| Bernalillo | 6.3 | 51,643 | 71,513 | 60.1 | 42.3 | 9.2 | 16.5 |
| Pima | 7.6 | 51,037 | 70,610 | 62.1 | 41.5 | 9.9 | 16.2 |
| Fresno | 9.4 | 51,261 | 71,663 | 51.7 | 48.7 | 9.5 | 21.3 |
| (Maricopa) | — | — | — | — | — | — | — |
| Sacramento | 7.4 | 63,902 | 85,289 | 65.3 | 39.8 | 6.3 | 14.3 |
| Fulton | 6.5 | 64,787 | 106,342 | 71.6 | 25.5 | 11 | 13.5 |
| Jackson | 5.1 | 52,805 | 71,871 | 69 | 30.2 | 11.6 | 13 |
| El Paso | 6.2 | 65,334 | 84,646 | 71.7 | 34.2 | 7.4 | 9.9 |
| Douglas | 4.2 | 61,688 | 84,908 | 73.4 | 26.2 | 9.6 | 7.2 |
| Wake | 4.3 | 76,956 | 102,190 | 78 | 22.2 | 8.9 | 8.4 |
| Miami-Dade | 6.3 | 48,982 | 75,744 | 72.3 | 6.4 | 22.5 | 16 |
| (Los Angeles) | — | — | — | — | — | — | — |
| Virginia Beach city | 4.9 | 74,186 | 93,675 | 80.4 | 24.9 | 8.2 | 7.5 |
| Alameda | 5.1 | 92,574 | 122,575 | 73.4 | 30.4 | 5.1 | 9 |
| Hennepin | 4.2 | 74,113 | 105,791 | 76 | 29.9 | 5.2 | 10.3 |
| Tulsa | 5.5 | 53,902 | 77,047 | 65.7 | 31 | 14 | 14.1 |
| Hillsborough | 6.1 | 56,137 | 79,843 | 63.5 | 32.3 | 13 | 14.7 |
| (Tarrant) | — | — | — | — | — | — | — |
| Orleans Parish | 8.4 | 39,576 | 69,101 | 55.8 | 41.6 | 10.7 | 23.8 |

## Results

Results of the meta-regression were summarized in Tables 2. A coefficient (slope of the meta-regression line) for COVID-19 prevalence was significantly negative for male sex (P < 0.001; Figure 1, Panel A), education attainment (P = 0.011), computer (P < 0.001; Figure 1, Panel B) and Internet (P < 0.001) use, and private health insurance (P = 0.029), which suggests that COVID-19 prevalence may decrease significantly as male sex, education attainment, computer and Internet use, and private health insurance increases. Whereas, the coefficient was significantly positive for black race (P < 0. 001), never matrimony (P < 0.001; Figure 1, Panel C), unemployment (P = 0.003), and poverty (P < 0.001), which suggests that COVID-19 prevalence may increase significantly as black race, never matrimony, and poverty increases. In the multivariable model entering all these 9 covariates, the coefficient was significantly negative for male sex (P = 0.036) and computer use (P = 0.024), and significantly positive for never matrimony (P < 0.001), which suggests that male sex and computer use may be independently and negatively associated with COVID-19 prevalence and never matrimony may be independently and positively associated with COVID-19 prevalence.

**Table 2.**
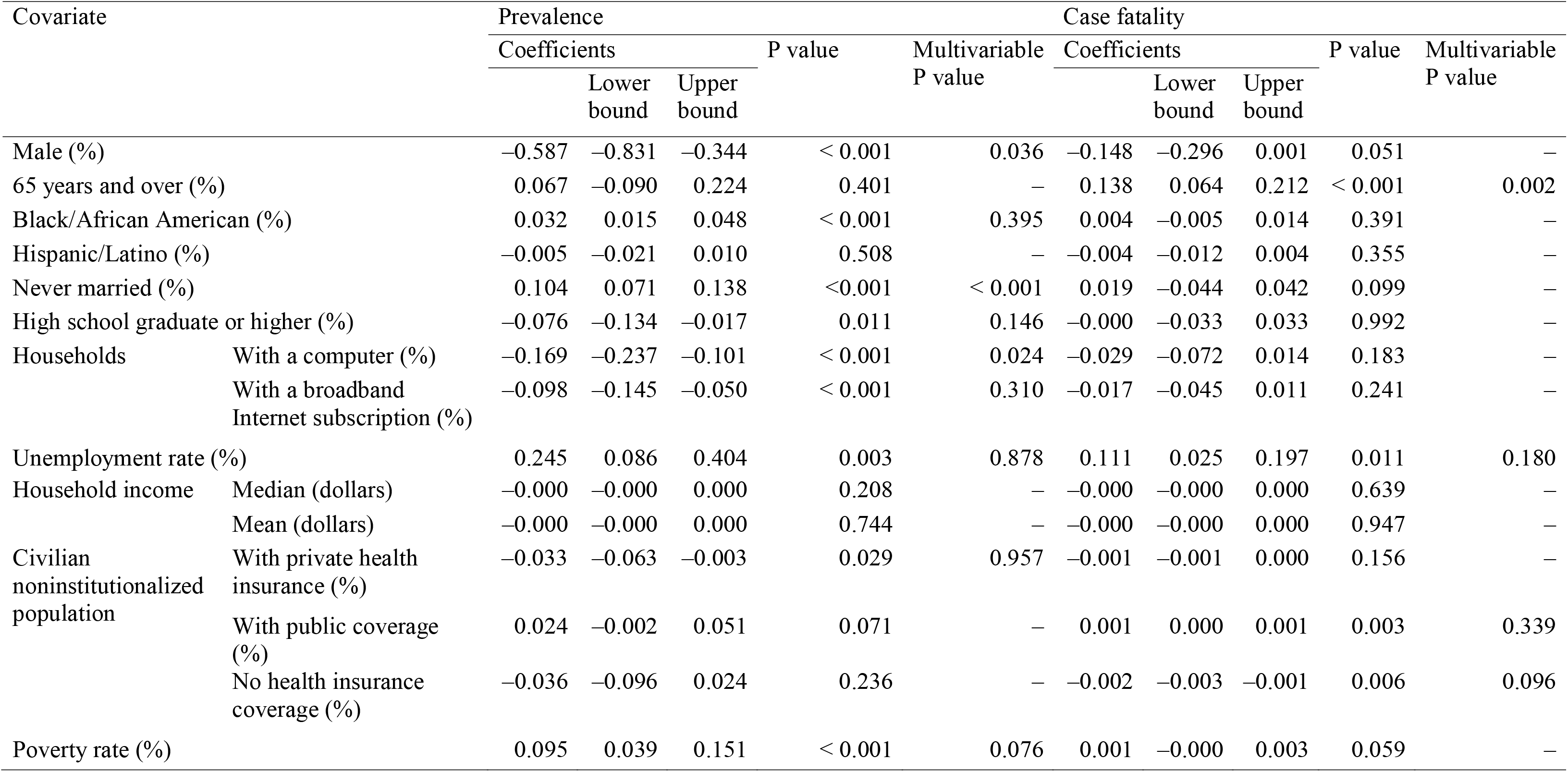
Meta-regression summary

**Figure 1.**
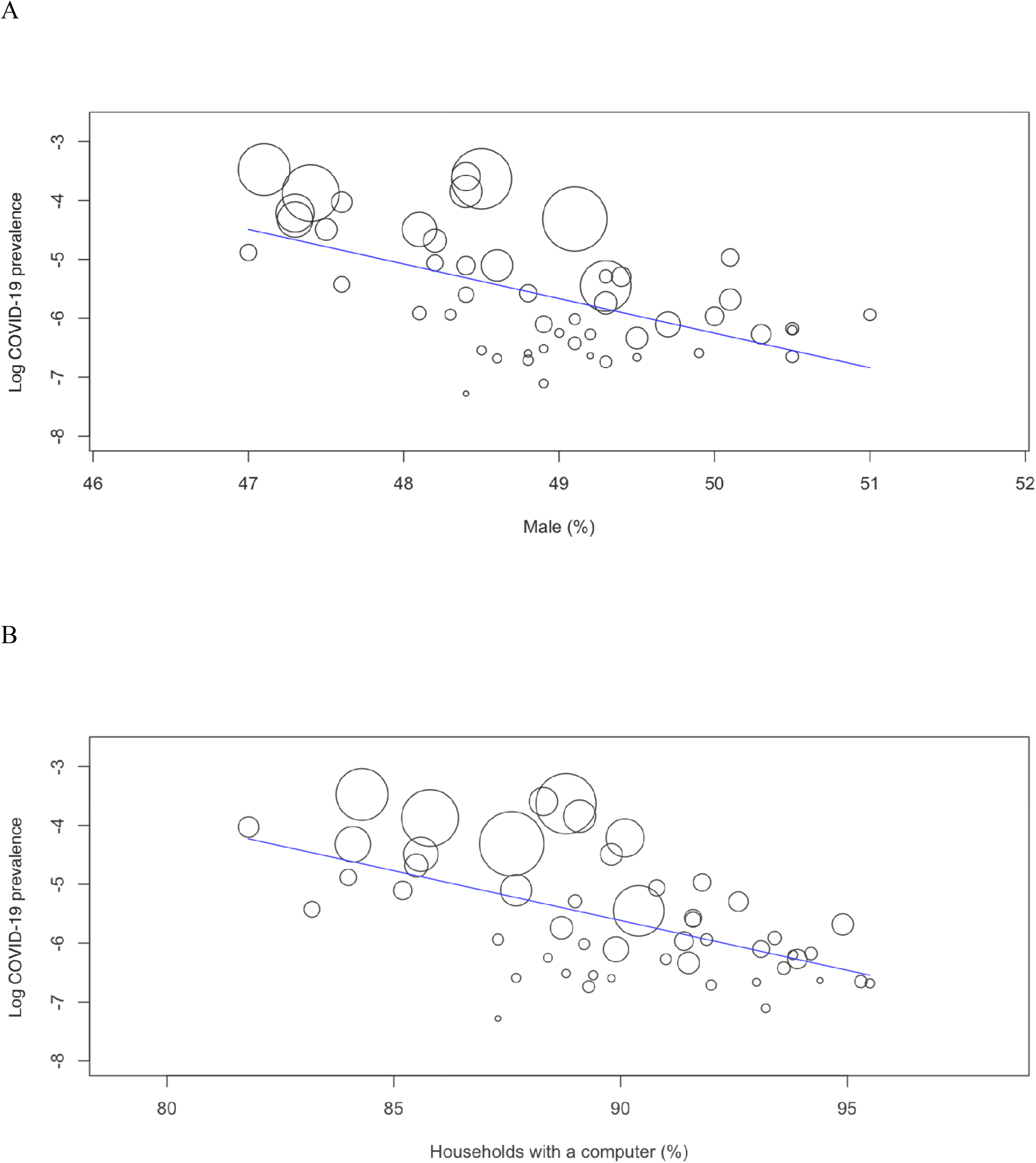
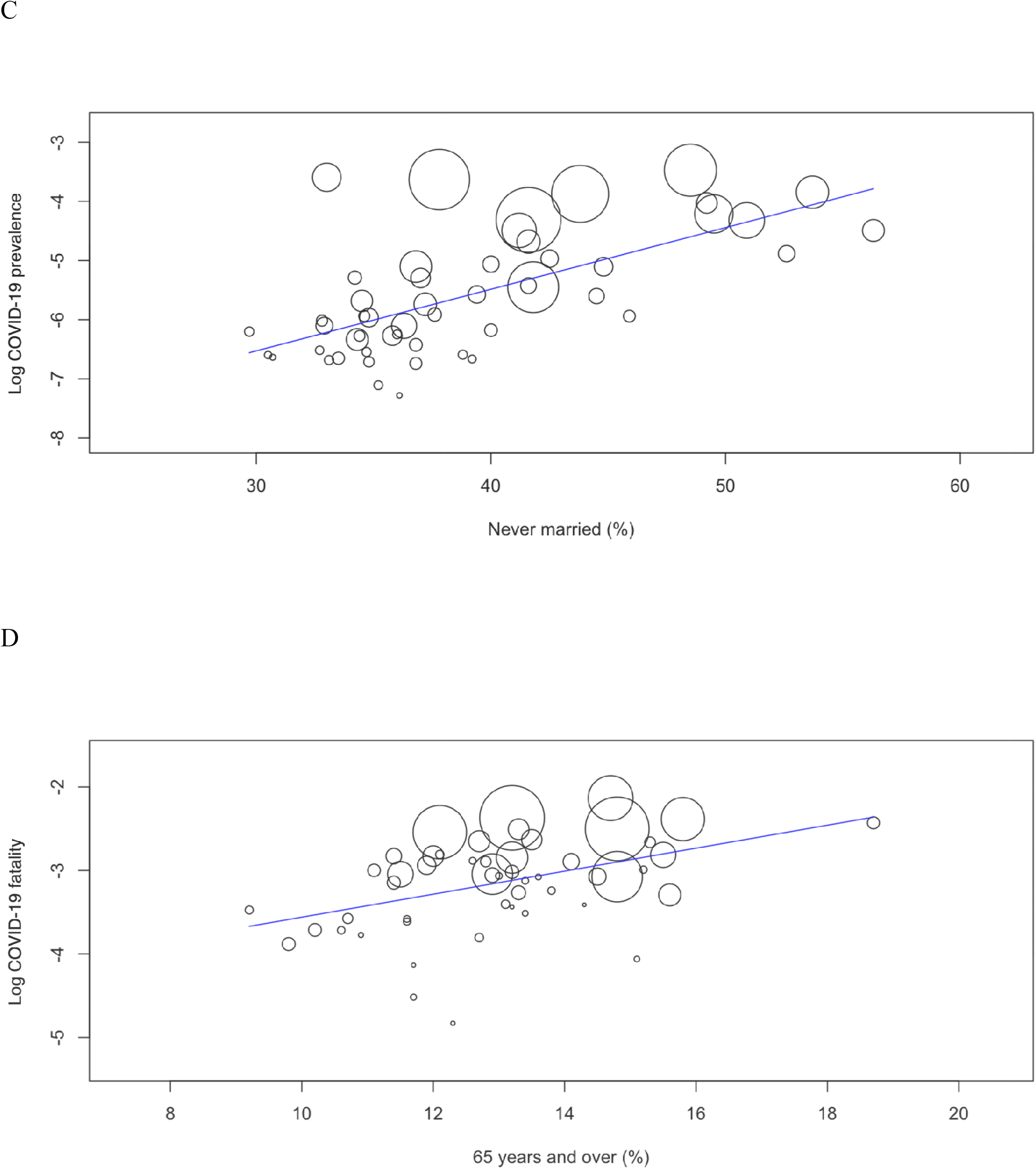
Meta-regression graph depicting the COVID-19 prevalence (Panels A–C) or fatality (Panel D) (plotted as the logarithm-transformed prevalence or fatality on the y-axis) as a function of a factor (plotted as a socioeconomic characteristic on the x-axis).

A coefficient for COVID-19 fatality was significantly negative for no health insurance (P = 0.006), and significantly positive for elderly (P < 0.001; Figure 1, Panel D), unemployment (P = 0.011), and public coverage (P = 0.003). In the multivariable model, the coefficient was significantly positive for only elderly (P = 0.002), which suggests that elderly may be independently and positively associated with COVID-19 fatality.

## Discussion

The present results suggest an independent and negative association of male sex and computer use with COVID-19 prevalence, an independent and positive association of never matrimony with COVID-19 prevalence, and an independent and positive association of elderly with COVID-19 fatality.

The present findings never denote, for instance, that a subject having a computer is at low risk of COVID-19 prevalence, which should be noted. Because of the community-level epidemiological study, the present results simply denote, for instance, that COVID-19 prevalence is lower in a community in which there are more subjects having a computer. Further patient-level clinical studies would be expected to determine, for instance, whether a subject having a computer is at low risk of COVID-19 prevalence. In meta-regression being generally different from simple regression, the relationship between outcome and explanatory variables is more influenced by larger studies (countries in case of the present analysis) than by smaller studies because the precision of each estimate weights studies.^3^ Thus, to screen potential risk and protective socioeconomic factors for COVID-19 prevalence and fatality, meta-regression may be more valid than simple regression.

In conclusion, a number of socioeconomic factors, e.g. male sex (negatively for prevalence), elderly (positively for fatality), never matrimony (positively for prevalence), and computer use (negatively for prevalence) may be associated with COVID-19.

## Data Availability

The datasets generated during and/or analysed during the current study are available from the corresponding author on reasonable request.

